# Monopolar network mapping predicts tremor outcomes of deep brain stimulation in Parkinson’s disease

**DOI:** 10.1101/2025.11.18.25340336

**Authors:** Alexander Calvano, Urs Kleinholdermann, Armin Kahn, Kenan Steidel, Felix Zahnert, Marina C. Ruppert-Junck, Philipp A. Loehrer, Miriam H.A. Bopp, Christopher Nimsky, Lars Timmermann, David J. Pedrosa

## Abstract

Deep brain stimulation is an effective therapy for tremor in Parkinson’s disease, yet clinical outcomes vary substantially across individuals. This variability highlights the need for continuous and quantitative measures that more accurately capture symptom severity and treatment response. Although precise subthalamic targeting remains fundamental to effective therapy, accounting for network-level modulation of disrupted motor circuits underlying core symptoms may further enhance clinical benefit. Nevertheless, the relative contributions of local and network-level effects to tremor suppression remain unclear, limiting our understanding of the mechanisms driving clinical improvement.

In this study, we identified a patient-specific connectivity fingerprint associated with effective tremor suppression following subthalamic deep brain stimulation. Twenty patients with Parkinson’s disease underwent objective tremor quantification using accelerometry during systematic monopolar reviews. For each stimulation setting, the corresponding volume of tissue activated served as a seed for probabilistic tractography, informed by individual diffusion-weighted imaging and subject-specific parcellation of grey and white matter structures. Subsequently, partial least squares regression was employed to analyse over 1,500 stimulation conditions, aiming to identify brain regions whose connectivity patterns predicted tremor outcomes. Ultimately, the resulting network model was compared with a local-effects model to assess whether network mapping provided superior predictive capability for motor improvement compared to traditional local targeting methods.

Our results demonstrate that (1) modulation of the basal ganglia and the cerebello-thalamo-cortical circuit at the network level predicts clinical improvement more accurately than local stimulation alone (*P* < 0.001); and (2) optimal stimulation sites for tremor relief are located in white-matter regions adjacent to the subthalamic nucleus. Model significance was confirmed through permutation testing (*P* < 0.001).

Based on these findings, we propose a framework that combines sensor-based measurements with monopolar mapping to investigate the effects of symptom-specific neuromodulation and to guide the selection of connectomic parameters for personalised treatment of motor symptoms.

## Introduction

Tremor is a debilitating motor symptom in Parkinson’s disease, which affects up to two-thirds of patients during disease progression. It often necessitates interventional therapy for adequate symptom control.^1,2^ While its severity is typically evaluated using clinical rating scales, motor fluctuations, short observation periods and insufficient inter-rater reliability may hamper these assessments.^3^ Mobile sensors offer a practical solution for continuous monitoring of objective motor impairment, allowing for accurate characterisation of temporal tremor patterns.^4,5^

Unlike other cardinal symptoms in Parkinson’s disease, the aberrant synchronisation underlying tremor is thought to arise from a complex pathological interaction between basal ganglia and cerebello-thalamo-cortical circuits.^6^ Although subthalamic deep brain stimulation (STN-DBS) is highly effective in mitigating tremor^7^, the mechanisms responsible for these effects remain elusive.^8^ Clinical programming has traditionally focused on applying current to the dorsolateral motor subregion of the STN^9,10^, with monopolar review remaining the standard approach for optimal contact selection.^11^ Recently, however, a growing body of evidence indicates that DBS leverages its effects not only from direct stimulation of key basal ganglia structures, but also from modulation of anatomically connected motor circuits and large-scale brain networks.^12^ Nevertheless, a direct comparison between local and network-level effects on objective tremor modulation has yet to be undertaken. Clarifying their respective contributions to clinical outcomes may enhance future programming strategies and decrease the proportion of suboptimal responders who continue to experience tremor despite precise lead placement.^13,14^

From a network perspective of movement disorders, connectivity profiles derived from local stimulation volumes have become valuable tools for identifying the anatomical substrates of specific symptoms and forecasting neuromodulatory effects.^12,15–17^ Recent studies have effectively combined ordinal ratings from monopolar review with normative connectomes to identify connectivity fingerprints of therapeutic and adverse effects.^12,18^ While this approach has provided valuable insights, normative atlases may fail to explain interindividual variability^15,19^ and reliance on ordinal data rather than continuous measurements further limits the precision of symptom-network associations.^20,21^

To address these limitations, we utilised preoperative diffusion-weighted imaging in 20 well-characterised patients with Parkinson’s disease to create a patient-specific connectivity fingerprint that reflects changes in objective tremor burden assessed through accelerometry. Notably, tremor modulation was quantified during monopolar review, allowing for a comprehensive evaluation of stimulation effects across individual contacts. We applied dimensionality reduction and regression analyses to evaluate the predictive utility of these connectivity fingerprints, comparing their performance directly against a model based on local effects. This approach allowed us to determine whether network mapping outperforms traditional anatomical targeting in predicting objective motor outcomes.

## Materials and methods

### Patients

Twenty tremor-dominant or equivalent-type patients with Parkinson’s disease (six females; 60.2 ± 6.9 years) from the Department of Neurology at the Philipps-University Marburg were included in the study. All participants had undergone chronic STN-DBS electrode surgery (Boston Scientific Neuromodulation Corporation, Valencia, CA 91355, USA) and declared written informed consent before participation. The study was approved by the local ethics committee (reference: 21/19) and adhered to the principles of the latest version of the Declaration of Helsinki. For descriptive analyses, see Table 1.

**Table 1:** Clinical details.

| ID | DD<br>[years] | MDS-UDPRS<br>III<br>(Med OFF,<br>Stim OFF) | MDS-UDPRS<br>III<br>(Med ON,<br>Stim ON) | Subtype<sup>a</sup> | LEDD<br>[mg] | Side<br>prevalence<sup>b</sup> |
| --- | --- | --- | --- | --- | --- | --- |
| 1 | 8 | 31 | 14 | equivalent-type | 835 | left |
| 2 | 5 | 36 | 23 | tremor-dominant | 550 | right |
| 3 | 4 | 39 | 30 | equivalent-type | 400 | right |
| 4 | 12 | 20 | 13 | tremor-dominant | 120 | right |
| 5 | 18 | 35 | 19 | equivalent-type | 895 | right |
| 6 | 13 | 69 | 21 | equivalent-type | 0 | left |
| 7 | 12 | 42 | 30 | equivalent-type | 525 | right |
| 8 | 8 | 33 | 9 | tremor-dominant | 0 | left |
| 9 | 4 | 40 | 16 | tremor-dominant | 200 | right |
| 10 | 3 | 44 | 14 | equivalent-type | 350 | left |
| 11 | 12 | 39 | 5 | equivalent-type | 225 | left |
| 12 | 12 | 26 | 5 | equivalent-type | 825 | right |
| 13 | 11 | 50 | 13 | equivalent-type | 732 | right |
| 14 | 2 | 64 | 24 | tremor-dominant | 105 | left |
| 15 | 8 | 56 | 23 | equivalent-type | 785 | left |
| 16 | 10 | 14 | 5 | equivalent-type | 595 | left |
| 17 | 9 | 41 | 21 | equivalent-type | 785 | right |
| 18 | 14 | 42 | 11 | tremor-dominant | 665 | right |
| 19 | 3 | 25 | 7 | equivalent-type | 277 | right |
| 20 | 13 | 56 | 29 | equivalent-type | 100 | right |
| Mean ±<br>SD | 9.1 ± 4.4 | 40.1 ± 14.0 | 16.6 ± 8.3 | NA | 448.5 ±<br>306.5 | NA |

Abbreviations: DD: Disease duration, LEDD: Levodopa equivalent daily dose, MDS-UPDRS: Movement Disorder Society-Unified Parkinson’s Disease Rating Scale, NA: Not applicable, SD: Standard deviation.
<sup>a</sup>Tremor-dominant: *n* = 6; equivalent-type: *n* = 14.
<sup>b</sup>Left-dominant: *n* = 8; right-dominant: *n* = 12.

### Clinical assessment

An overview of the methodological pipeline is shown in Fig. 1. Objective motor performance was evaluated three months after surgery in the OFF-medication state. Tremor was assessed using a commercially available wearable device, the Myo Gesture Control Armband (Thalmic Labs, Kitchener-Waterloo, Ontario). This device recorded angular velocity and linear acceleration through a gyroscope and accelerometer at a sampling rate of 50 Hz^4^, as described by Kleinholdermann *et* al.^22^ In brief, each of the 16 electrode contacts was tested in monopolar configuration, with stimulation amplitude increased in 0.5 mA steps up to a maximum of 5 mA or until intolerable side effects occurred. Stimulation frequency and pulse duration were set at 130 Hz and 60 µs, respectively. Four motor tasks (finger tapping, rotational hand movements, postural hand holding and rest posture) were evaluated separately for the left and right side at each amplitude level (Fig. 2). We regarded the resting task to be the most representative measure for analysing the relationship between different DBS settings and tremor outcomes. This was based on two considerations: (i) the high sensitivity of mobile sensors in detecting subtle changes in motor symptoms^23^; and (ii) the fact that not all tremor-dominant or equivalent-type Parkinson’s disease patients exhibit postural tremor.^24^ To this end, root mean square (RMS) scores were derived from accelerometer signals across each trial. Raw data were demeaned, linearly detrended and band-pass filtered with a 4th-order Butterworth filter (2–15 Hz). The resulting RMS scores were log-transformed to normalise their distribution for analysis.

**Figure 1.**
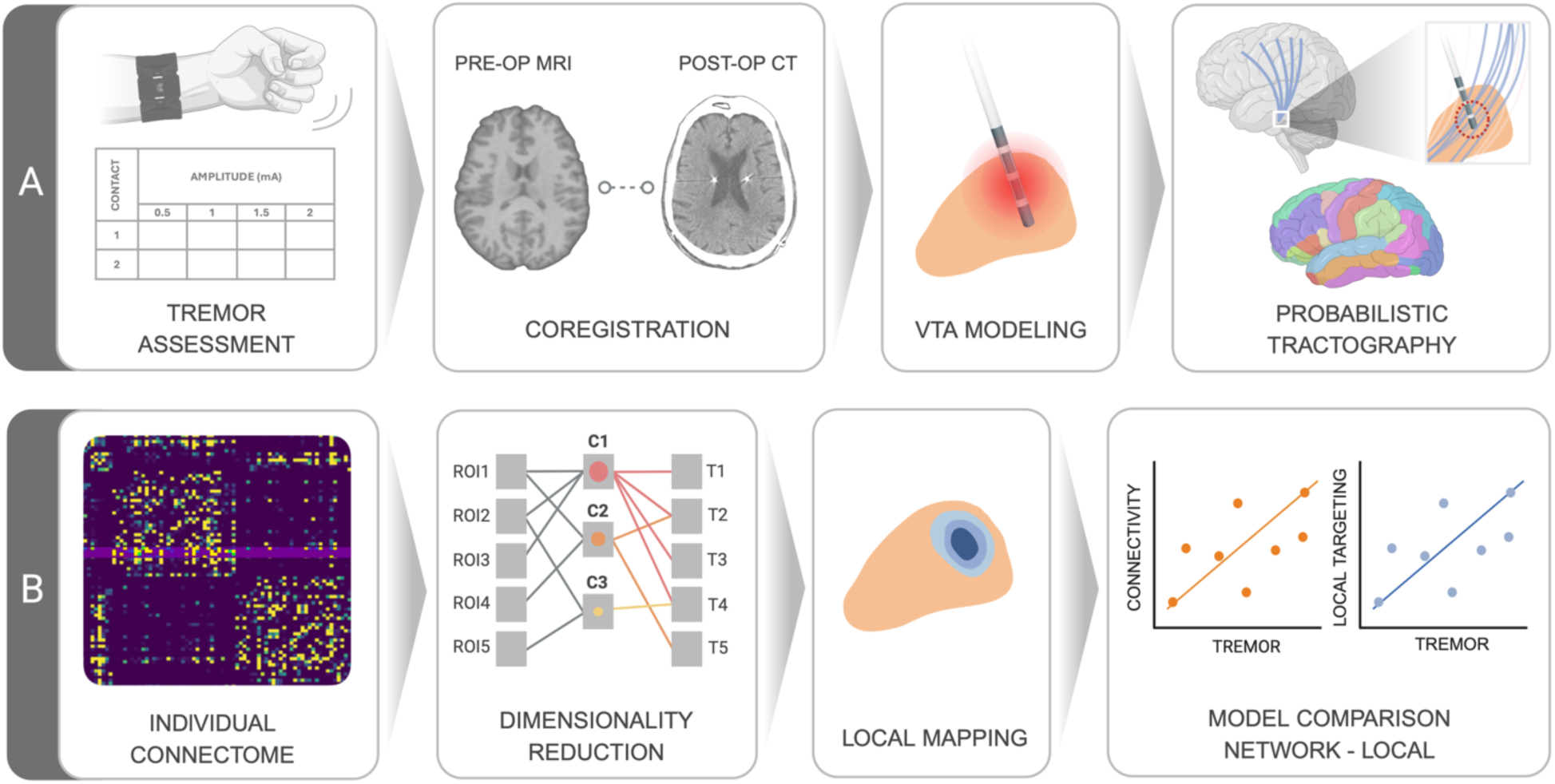
Methodological pipeline. (**A**) Tremor severity was quantified during systematic review of monopolar contacts using accelerometry. Image processing included coregistration of preoperative MRI with postoperative CT scans, followed by modelling the volumes of tissue activated (VTA) for each current intensity and contact level. Each VTA served as a seed region for probabilistic tractography to generate individual structural connectomes. (**B**) Fibre counts from VTAs to the connectome were analysed using partial least squares (PLS) regression. The resulting network model was compared with a local effects model to assess predictive utility. Created in BioRender (https://BioRender.com/d1o347l).

**Figure 2.**
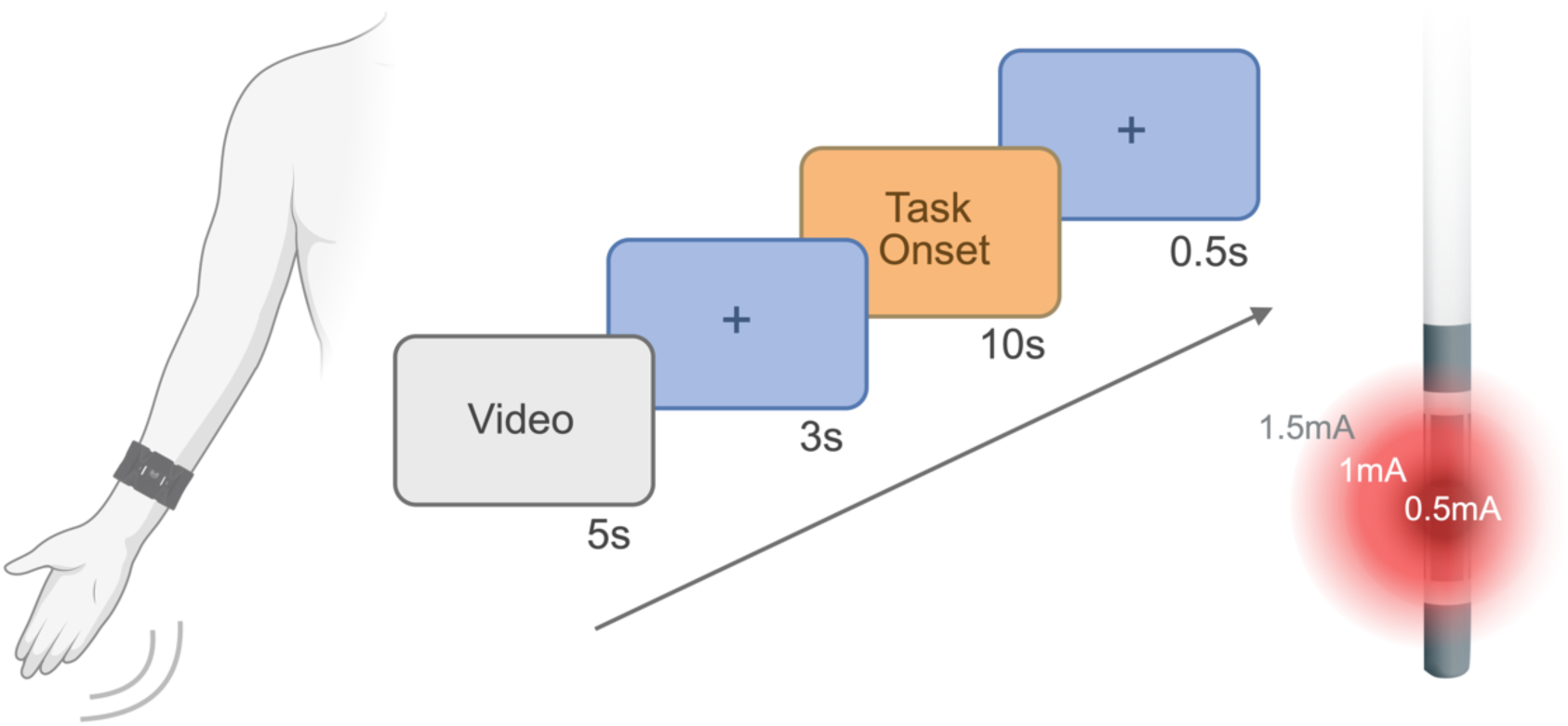
Motor paradigm across stimulation amplitudes and contacts. Participants performed four simple motor tasks (finger tapping, rotational hand movements, postural hand holding and rest posture) during sequential monopolar activation of contacts (c1–c8, c9–c16) at increasing current intensities for both sides. Motor burden was assessed using wearable motion sensors. Created in BioRender (https://BioRender.com/9byytrw).

### Modelling of stimulation volumes

As changes in motor performance can arise from stimulation beyond predefined target structures and across different current intensities, volumes of tissue activated (VTAs) were modeled for every tested stimulation amplitude in each patient according to the Lead-DBS pipeline (version 3.1) (https://www.lead-dbs.org).^25^ Briefly, after coregistration of preoperative MRI and postoperative CT scans, all images were spatially normalised into the standard MNI152 space.^26^ Leads were localised using the PaCER algorithm.^27^ A finite element model was employed to compute stimulation volumes by estimating the gradient distribution of the electrical charge in space on a tetrahedral mesh that differentiated four compartments: grey matter, white matter, electrode contact and insulation.^19^ An electric field threshold of 0.2 V/mm was used during estimation.^28,29^ Subsequently, a linear rigid-body transformation was applied to register VTAs from individual structural space to diffusion space, as implemented in SPM (version 12).^30^

### Image processing

For all but four participants, preoperative MRI was acquired with a 3 T MRI system (Tim Trio, Siemens Healthineers, Erlangen, Germany).^31^ The remaining patients were scanned on a 1.5 T MRI (Siemens Avanto, Erlangen, Germany) during clinical routine. The imaging protocol for all participants is included in the Supplementary material.

Processing of structural MRI was performed with the recon-all pipeline in FreeSurfer (version 7.4.1) (http://surfer.nmr.mgh.harvard.edu). The processing of T1-weighted scans included skull stripping, automated Talairach transformation, cortical and subcortical segmentation, intensity normalisation, tessellation of the grey/white matter boundary and automated topology correction.^31,32^ The Desikan-Killiany parcellation^33^ was used for the definition of regions of interest (ROIs), which were registered from individual structural space to the diffusion-weighted image via boundary-based registration.^34^

Diffusion-weighted images were processed with the FMRIB Software Library (FSL) 6.0.7.15 (https://fsl.fmrib.ox.ac.uk/fsl). Eddy-current distortions and head motion were corrected through linear registration and resampling of raw diffusion-weighted images to the first b0 volume (FSL’s eddy).^35^ Fibre orientations were then estimated with BedpostX, permitting for up to three crossing fibre populations in each voxel.^36^ Probabilistic tractography was conducted with the GPU version of ProbtrackX2 in network mode, with 30,000 streamlines seeded per voxel of each VTA-derived parcel.^36,37^ Tractography was confined to cortical voxels at the grey/white matter boundary, while the remaining cortical grey matter was defined as an exclusion mask to prevent spurious connections across adjacent gyri. Additional exclusion masks were specified for cerebrospinal fluid and ventricles, whereas the whole white matter served as a waypoint mask.^38^ For each stimulation level at every contact, this yielded an individual asymmetric 84 × 84 (ROI × ROI) connectivity matrix, where the elements represented the number of streamlines as a measure of connection strength. Streamline counts from bilateral VTAs to the rest of the connectome were extracted for downstream analyses.

### Local mapping

To assess stimulation outcomes at a local level, a clinical effectiveness map was generated based on all VTAs with corresponding tremor scores. To this end, each binarised VTA (*V_i_*) was multiplied voxelwise by the inverse of its contralateral RMS score (*R_i_*), thereby assigning greater weight to VTAs associated with better clinical outcomes. The effectiveness map (*E*) was then computed as the sum of these weighted images divided by the sum of the non-weighted images (1). VTA processing was performed at an isotropic resolution of 1 × 1 × 1 mm^3^.

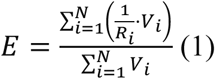

To account for differences in volume size, mean intensity values (MIV) were calculated by adding the voxel intensities (*x*) within each VTA (*V_k_*) in the effectiveness map (*E*) and dividing by the number of voxels in the respective VTA mask (|*V_k_*|) (2). Following this approach, each VTA was assigned a distinct MIV, which served as a quantitative surrogate of local stimulation response.

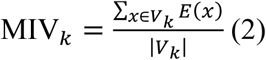

### Statistical analysis

With a partial least squares (PLS) regression, we evaluated the relationship between structural connectivity and objective tremor performance using the pls package in R (version 2.8-1). PLS identifies latent components that maximise the shared variance between two datasets.^39^ A ten-fold cross validation was conducted using the package caret (version 7.0-1) for model training and out-of-fold predictions were collected to assess model performance. After optimisation through model training, six components were retained, with component loadings reflecting the relative contribution of each VTA-ROI connection to tremor reduction. Model significance was assessed using permutation testing with 1,000 simulations of randomly shuffled outcome values. Wilcoxon signed-rank tests were performed to compare RMS scores and VTA volumes between hemispheres, with results expressed as median and interquartile ranges (Q1–Q3). Furthermore, associations between MIVs and tremor scores were assessed using Spearman correlation, while Pearson correlation was used to evaluate the relationship between true and predicted values in the network and local effects model. Both models were compared using Steiger’s Z test. Scripts for data processing and visualisation were written in MATLAB (version R2023b), Bash and Python (version 3.12.3) using the packages nibabel, numpy, pandas and matplotlib. Glass brain visualisations were created with the Python package nilearn (version 0.11.1) (https://nilearn.github.io/stable/index.html) and statistical analyses were conducted in R (version 4.3.0).^40^ Statistical significance was defined as *P* < 0.05.

## Results

The location of all DBS leads in MNI152 space is shown in Fig. 3A. In total, 1,561 VTAs with matching tremor scores were computed (left = 804, right = 757). Volume sizes did not differ between hemispheres (left: 21 mm³ [8–49], right: 20 mm³ [8–43]; *P* = 0.13) (Fig. 3B). Similarly, stimulation amplitudes were identical for both sides (left: 1.0 mA [0–2.5], right: 1.0 mA [0–2.5]). Across all tested stimulation settings, RMS scores indicated a greater motor affection on the right (9.9 [5.7–20.7]) compared to the left side (7.4 [5.3–15.4]; *P* < 0.001) (Fig. 3C).

**Figure 3.**
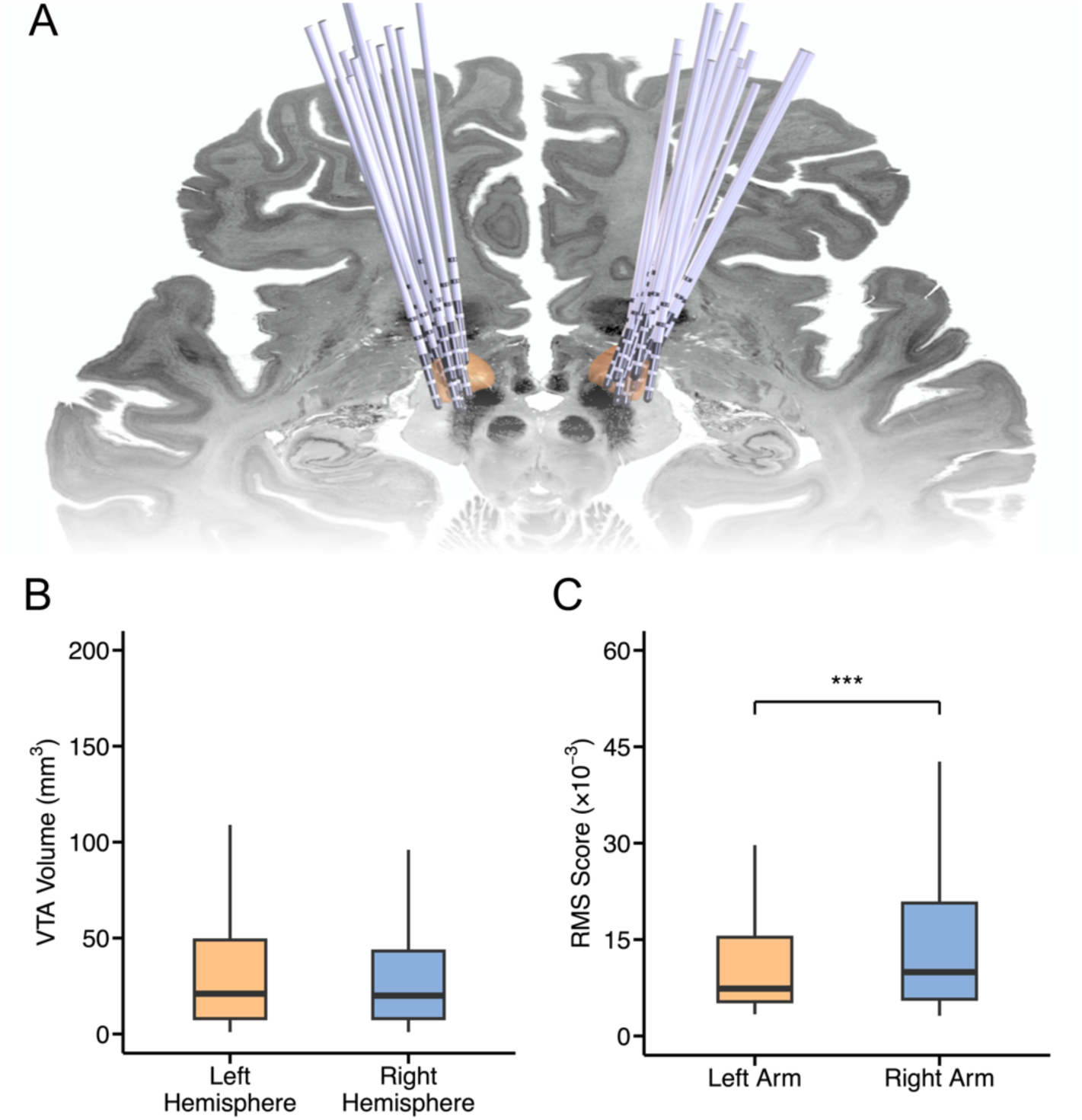
Lead locations and tremor outcomes. (**A**) Electrode positions of all participants in MNI152 space. The subthalamic nucleus (STN) is illustrated in brown; parcellation according to the DISTAL minimal atlas.^68^ (**B–C**) Boxplots show comparisons between stimulation volumes and root mean square (RMS) scores between left and right. Central marks indicate the median and edges the 25th and 75th percentiles of the distribution. \*\*\**P* < 0.001 (Wilcoxon signed-rank test).

### Network model

For the left hemisphere, the six retained components accounted for 57% of the predictor variance (network connectivity) and 37% of the variance in tremor outcomes. In the right hemisphere, they accounted for 61% and 47% of the predictor and outcome variances, respectively. For better overview, only the top three components explaining the largest proportion of both datasets are described here. The first component (C1) (left: 38% / 5%, right: 14% / 16%) was characterised by loadings reflecting structural connections between VTAs and the pallidum, putamen, caudate, thalamus, cerebellum and the pre- and postcentral gyrus. The third component (C3) (left: 6% / 6%, right: 6% / 11%) demonstrated a highly similar pattern, with stronger loadings for the right hemisphere. Together, these findings suggest that tremor suppression is predominantly mediated through cerebello-thalamo-cortical and basal ganglia-thalamo-cortical networks. In contrast, the second component (C2) (left: 5% / 16%, right: 33 % / 6%) was defined by loadings in associative and paralimbic regions, including the middle temporal gyrus, rostral middle frontal gyrus, inferior parietal lobule, frontal pole, supramarginal gyrus, lateral orbitofrontal cortex and hippocampus, reflecting a distinct non-motor network profile (Fig. 4). All components with their respective network loadings are provided in the Supplementary material (Supplementary Tables 1–6). The obtained network model was associated with tremor scores in both hemispheres (left: *r* = 0.51, right: *r* = 0.61; both *P* < 0.001). Permutation testing (1,000 iterations) demonstrated that the observed model performance was unlikely to occur by chance (*P* < 0.001).

**Figure 4.**
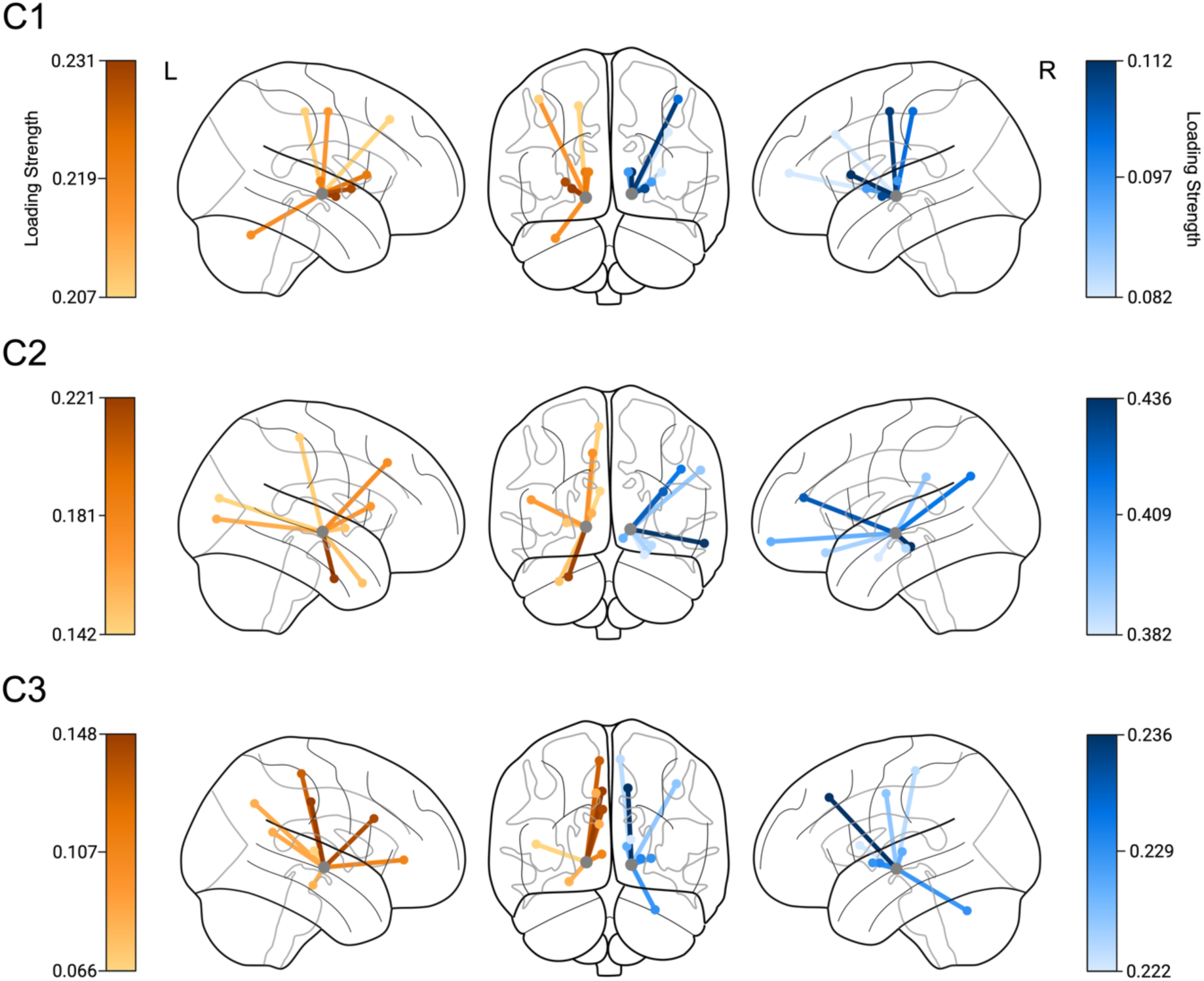
Component loadings of structural connectivity. Glass brains depict structural connections in MNI152 space between volumes of tissue activated (VTA, grey) to regions of interest (ROIs) associated with tremor outcomes for the left (orange) and right (blue) hemispheres; parcellation according to the Desikan-Killiany atlas.^33^ Node colours indicate loading strength derived from partial least squares (PLS) analysis. Components 1 and 3 converge on a common network involving the basal ganglia, thalamus, cerebellum and motor cortex. In contrast, component 2 highlights connections with associative and paralimbic regions.

### Local effects

All VTAs with corresponding tremor scores were used to generate clinical effectiveness maps for each hemisphere. The resulting volume was slightly larger for the right hemisphere (1,437 mm^3^) compared to the left hemisphere (1,352 mm^3^). Visual inspection of the effectiveness maps indicated that peak intensities were not confined to the STN itself, but instead seemed to be located dorsolateral to the nucleus. The most effective clusters in both hemispheres appeared to be situated in lateral white matter regions adjacent to the STN and close to the caudal Zona incerta (cZi) (Fig. 5). To demonstrate the degree of fit between the model and tremor data, MIVs were correlated with contralateral RMS scores for both hemispheres (left: rho = –0.51, right: rho = –0.35; both *P* < 0.001) (Supplementary Fig. 1). As these correlations are somewhat circular, given that the effectiveness maps were derived from tremor scores, we refrained from reporting *P*-values of these associations.

**Figure 5.**
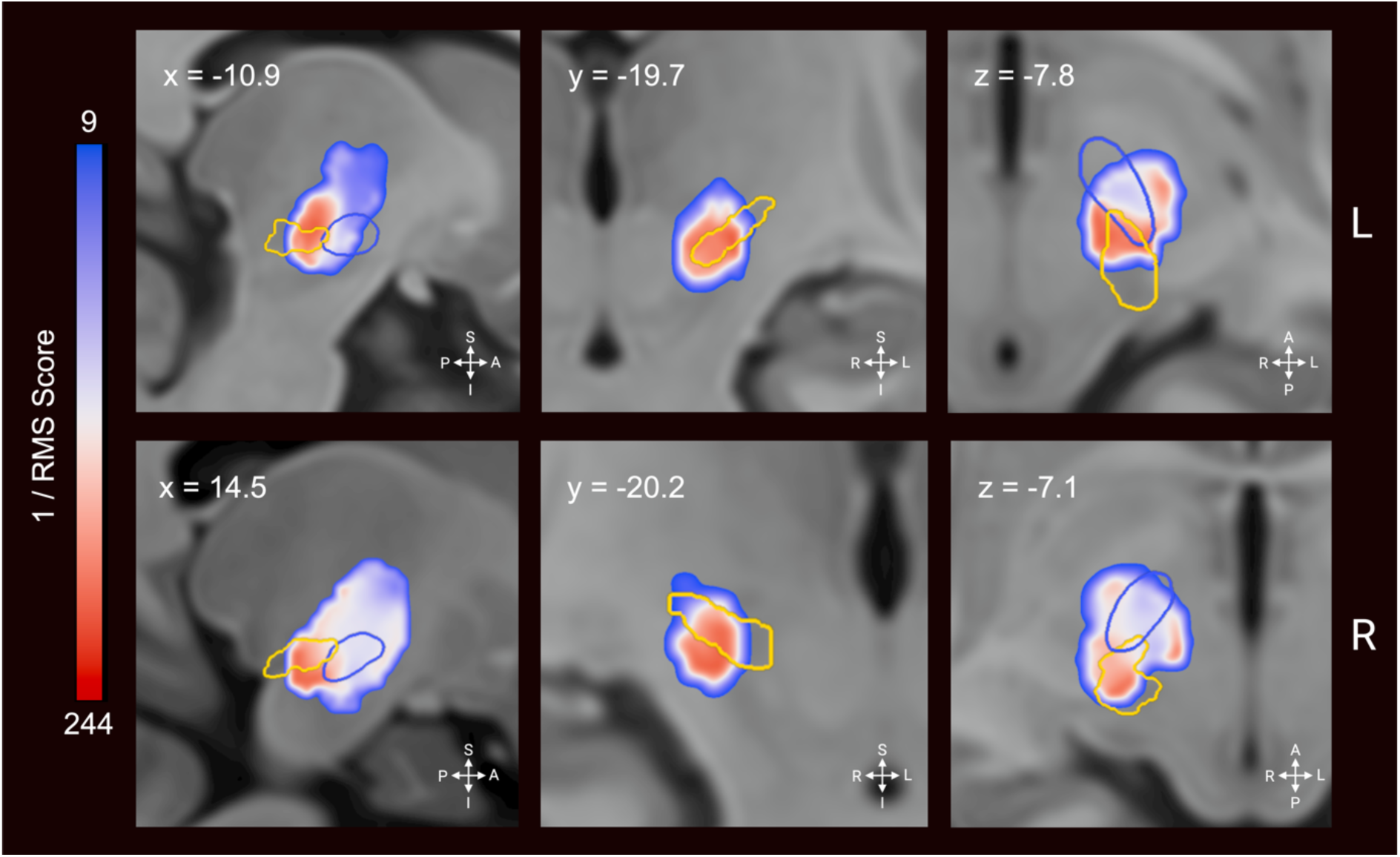
Spatial distribution of stimulation effects. Clinical effectiveness maps are shown in sagittal (x), coronal (y) and axial (z) views for the left (top row) and right hemisphere (bottom row) in MNI152 space. Outlines denote the subthalamic nucleus (STN, blue) and caudal zona incerta (cZI, yellow); parcellation according to the DISTAL minimal atlas ^68^ and the Zona incerta atlas ^69^, respectively. Regions of greater tremor improvement (warmer colors) appear to predominantly reside in white matter regions and the caudal Zona incerta (cZi), with partial extension into the dorsolateral STN.

### Model comparison

To assess whether network features offer superior predictive performance compared to local stimulation effects, we compared the PLS model with the linear regression model reflecting local stimulation effects. For the left hemisphere, the PLS model (*R*^2^ = 0.26) predicted tremor outcomes better than the linear model (*R*^2^ = 0.08). For the right hemisphere, the difference was even more substantial (PLS: *R*^2^ = 0.37, linear model: *R*^2^ = 0.08) (Fig. 6). Steiger’s Z tests confirmed the superiority of the PLS approach for both hemispheres (left: *z* = –6.19, right: *z* = –8.92; both *P* < 0.001), indicating that network connectivity is more predictive of tremor outcomes than local stimulation alone.

**Figure 6.**
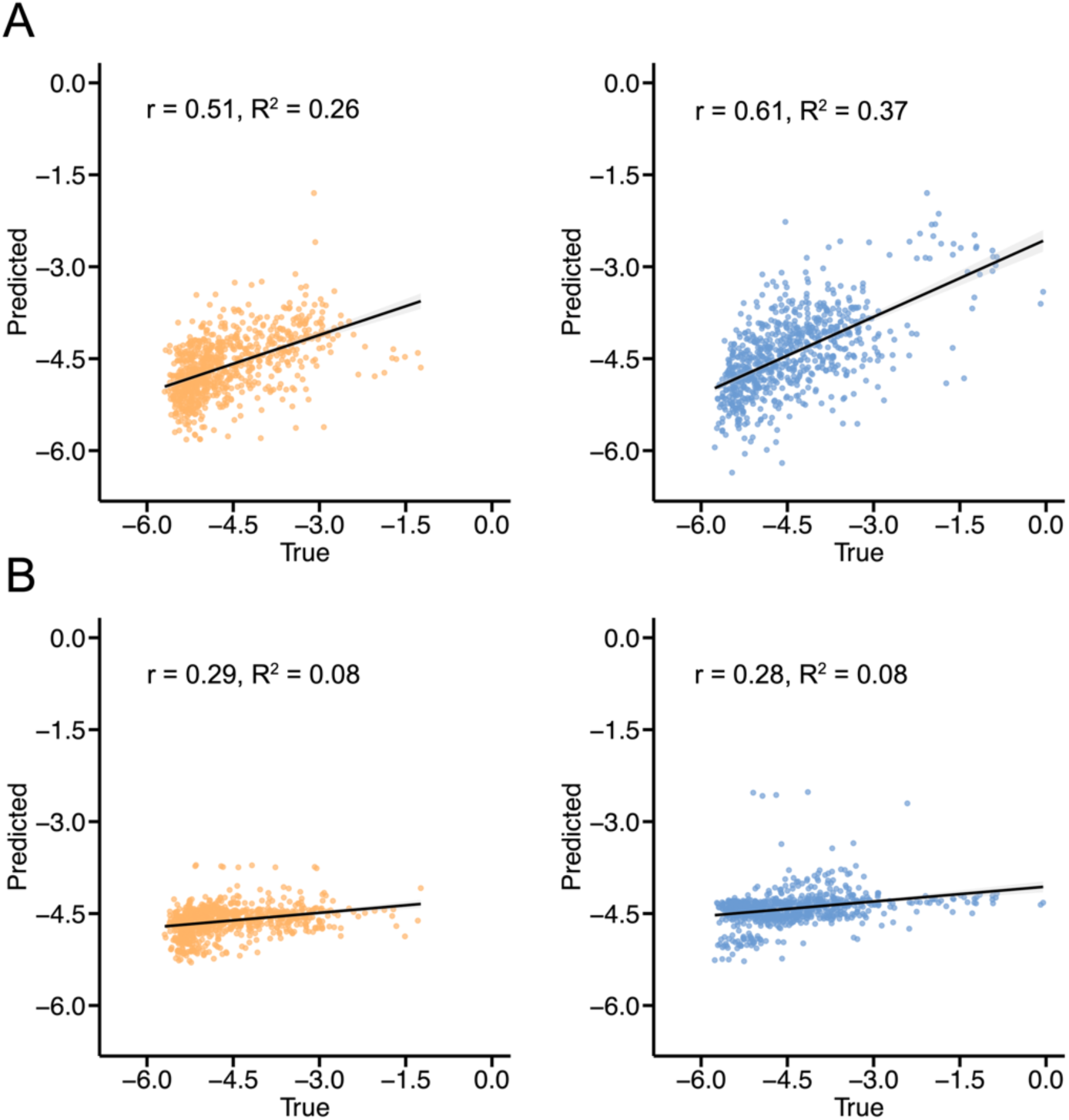
Comparison of model performance in predicting tremor outcomes. Scatter plots show Pearson correlations between true and predicted tremor scores for the network model (**A**) and the local effects model (**B**) across hemispheres (left = orange, right = blue). Black lines indicate linear regression fits.

## Discussion

In this study, we sought to delineate a structural connectivity profile for effective tremor suppression following STN-DBS and to test whether network-based models provide superior predictive power compared to local targeting. For the first time, objective tremor recordings obtained during monopolar review were used to construct a patient-specific connectivity model that integrated variations in spatial stimulation effects. Within this network, we observed distinct structural connections to the cerebello-thalamo-cortical circuit and basal ganglia structures to be particularly effective for tremor control. Crucially, we demonstrated that network level modulation outperformed the local effects model in clinical prediction, lending credence to the overarching idea of circuit level therapeutic effects of DBS.

The potential of DBS to restore pathological network dynamics at the macroscale has been demonstrated for several movement disorders.^29,41^ To delineate the spatial distribution of therapeutic pathways within given symptom domains, normative connectomes are frequently employed.^42^ Their large sample sizes confer high statistical power and facilitate the identification of consistent connectivity patterns across diverse populations.^19^ Nevertheless, a recent discourse has emerged concerning the relative specificity and sensitivity of tract-based versus normative connectomes as well as the potential contribution of histological data in reconciling their discrepancies.^43^ Furthermore, normative connectomes exhibit limitations in accounting for anatomical variability, particularly within the framework of neurodegeneration.^44,45^ This issue is particularly pertinent in the context of Parkinson’s disease, wherein distinct clinical subtypes demonstrate divergent patterns of structural and functional alterations.^46^ In a comparative study, Wang *et* al.^15^ showed that patient-specific and normative connectomes yield largely analogous optimal connectivity profiles for STN-DBS. However, patient-specific connectivity accounted for a greater proportion of variance in clinical improvement, indicating a potential advantage in capturing individual anatomical variability.

Prior studies investigating the anatomical correlates of effective tremor suppression have so far relied on ordinal rating scales^47–50^, which may be hampered by insufficient interrater and within-subject reliability.^51^ Furthermore, tremor severity is known to fluctuate substantially depending on motor state, medication status and external factors^52^, with daily amplitude variations of up to 23%.^53^ Consequently, reliance on these measurements carries the risk of distorting the extent of motor impairment^51^, thereby limiting the accuracy of identifying therapeutic targets. Translating symptom-network associations into clinical decision-making therefore not only necessitates patient-specific anatomical data, but also precise symptom monitoring for tailoring stimulation strategies to individual symptoms.^12,54^ To address these limitations, we integrated high-resolution, objective recordings of tremor severity with diffusion-weighted imaging and subject-specific parcellation of grey and white matter structures. In this setting, the resulting connectivity model demonstrated markedly higher predictive power for tremor outcomes than models based solely on local stimulation effects. Based on these findings, we propose a methodological framework that combines quantitative symptom measurement with monopolar mapping to study symptom-specific neuromodulation. This approach complements next generation DBS leads, which allow finer parameter control and may help clinicians target symptom-relevant fibre pathways while avoiding those linked to side effects.^41,55^

The concept that distinct network hubs account for specific symptoms in Parkinson’s disease is widely accepted.^6,41,56^ Our results provide further support for the cerebello-thalamo-cortical loop as a key anatomical substrate for tremor suppression.^57^ The relevance of this circuitry was recently corroborated by Goede *et* al.^47^, who identified an anatomical network in which tremor subtypes mutually unfold. Key regions included the primary motor cortex (M1), cerebellum, supplementary motor area and occipital regions, converging across structural, functional and lesion-based network maps. From a pathophysiological perspective, functional imaging from tremor-dominant Parkinson’s disease patients has shown that abnormal coupling between basal ganglia output structures and the cerebello-thalamo-cortical loop is associated specifically with tremor severity, but not with bradykinesia. Pallidal dopamine depletion appears to drive this pathological coupling, with transient increases in basal ganglia-cortical interactions triggering and sustaining tremor-frequency (∼6 Hz) oscillations within the network.^6,58^ By disrupting pallido-cortical synchronisation^59^, DBS of the internal globus pallidus may attenuate aberrant tremor signals by preventing downstream propagation to cerebellar connections.^47^ In STN-DBS, activation of the hyperdirect pathway is considered an important therapeutic mechanism.^60^ However, our results suggest that effective tremor control requires engagement of a broader network beyond the structural connections of the STN.

Our connectivity analysis highlights two structures as particularly important for tremor control: the dentatorubrothalamic tract (DRTT) and the cZi. The DRTT conveys cerebellar outflow from the dentate nucleus and ascends to the ventrolateral thalamus before projecting to cortical motor areas.^61^ Targeting this fibre bundle has been independently validated as an alternative DBS target for tremor of various origins, including essential tremor, dystonic tremor and Holmes tremor.^62^ Similarly, stimulation of the cZi yields greater tremor improvement on clinical scales than STN stimulation^63,64^ and robustly engages the hyperdirect pathway.^65^ Anatomically, the cZi lies between the Forel fields and near the pallidothalamic tract, which includes the lenticular fasciculus (Forel field H2) and the ansa lenticularis.^66^ Consistent with these findings, the highest signal intensities in our effectiveness maps were located in lateral white matter regions adjacent to the STN. Together, these results suggest that steering current toward the DRTT and cZi represents a promising strategy for tremor suppression in Parkinson’s disease, underscoring their role as integrative network hubs that can be selectively modulated using directional, patient-specific DBS.

### Limitations

Several limitations warrant acknowledgement in this study. Although most participants were scanned using identical MRI systems and acquisition protocols, four subjects underwent clinical diffusion-weighted MRI with different parameters. While previous work has demonstrated that such data can be successfully integrated into DBS connectivity analyses^21,44^, uniform protocols would have been preferable. The diffusion-weighted scans also had relatively low angular resolution, which limits the ability to resolve complex fibre crossings. In addition, it is important to emphasise that fibre counts remain an imperfect surrogate of connectivity strength and may not correspond to absolute biological values.^67^ A further methodological consideration in this context pertains to our selection of a tractography-based connectomic framework. In a recent study, Mehta *et* al.^43^ demonstrated marked inconsistencies in pathway activation profiles across tractography-based, normative and histology-based connectomes. Therefore, it is essential to interpret DBS network findings with caution, as the ability to predict clinical outcomes does not necessarily reflect anatomical accuracy. Another limitation concerns our use of incorporating multiple VTAs per contact for probabilistic tractography. This can introduce spatial dependencies, which may have inflated streamline counts due to overlapping stimulation volumes. Nevertheless, PLS analysis addresses this multicollinearity by reducing correlated connectivity profiles into orthogonal latent variables that maximise covariance with clinical outcomes.^39^ In other words, PLS analysis captured the most significant network structures while accounting for the fact that nearby stimulation sites share similar connections. Finally, it can be posited that the relatively small sample size may hamper the generalisability of our findings. However, we contend that the inclusion of over 1,500 stimulation settings has facilitated a robust sampling of objective stimulation-response relationships.

### Conclusion

Our findings demonstrate that effective tremor suppression arises primarily from network-level modulation involving the basal ganglia and cerebello-thalamo-cortical pathways, rather than from local stimulation alone. By systematically mapping monopolar contacts, we identified fibres adjacent to the subthalamic nucleus and embedded within these tremor-relevant circuits as the strongest predictors of clinical improvement. Together, these results highlight the utility of monopolar mapping for delineating symptom-specific networks and underscore the potential of patient-tailored fibre tracking to guide stimulation parameter selection and advance personalised, connectomic neuromodulation.

## Data availability

The programming code used for the analyses in this report is available in an online repository (https://github.com/AlexanderCalvano/tremorDBS). The data that support the findings of this study are available upon request from the corresponding author (AC). The raw data are not publicly available due to privacy or ethical restrictions.

## Acknowledgments

We would like to express our gratitude to all the patients who participated in our study.

## Funding

AC and FZ were supported by the Clinician Scientist Program of the Philipps-University Marburg. PAL was supported by the von Behring-Röntgen Stiftung and a Research Grant of the University Medical Centre Giessen and Marburg.

## Competing interests

AC has participated in a training course, which was industry funded by Stada Arzneimittel AG and received a travel grant from the Prof. Klaus Thiemann Foundation in the German Society of Neurology. LT reports grants, personal fees and non-financial support from SAPIENS Steering Brain Stimulation, Medtronic, Boston Scientific and St. Jude Medical and has received payments from Bayer Healthcare, UCB Schwarz Pharma and Archimedes Pharma and also honoraria as a speaker on symposia sponsored by Teva Pharma, Lundbeck Pharma, Bracco, Gianni PR, Medas Pharma, UCB Schwarz Pharma, Desitin Pharma, Boehringer Ingelheim, GSK, Eumecom, Orion Pharma, Medtronic, Boston Scientific, Cephalon, Abbott, GE Medical, Archimedes and Bayer. DP received honoraria as a speaker at symposia sponsored by Boston Scientific Corp, Medtronic, AbbVie Inc., Zambon and Esteve Pharmaceuticals GmbH. He received payments as a consultant for Boston Scientific Corp and Bayer and he received a scientific grant from Boston Scientific Corp for a project entitled: ‘Sensor-based optimisation of Deep Brain Stimulation settings in Parkinson’s disease’ (compareDBS). Finally, DP was reimbursed by Esteve Pharmaceuticals GmbH and Boston Scientific Corp for travel expenses to attend congresses. KS has participated in training courses by Boston Scientific Corp and St. Jude Medical and was reimbursed for travel costs for attending conferences by Boston Scientific Corp. FZ received funding from the LOEWE Research Cluster ADMIT (Advanced Medical Physics in Imaging and Therapy) and was further reimbursed for travel expenses by UCB. MB and CN are scientific consultants for Brainlab. The remaining authors declare that they have no known competing financial interests or personal relationships that could have appeared to influence the work reported in this paper.

## Notes

### Author Declarations

The ethics committee of the Philipps-University Marburg gave ethical approval for this work

