## Supplementary Material for "Monopolar network mapping predicts tremor outcomes of deep brain stimulation in Parkinson’s disease"

#### MRI data acquisition and processing

3 T MRI system (Tim Trio, Siemens Healthineers, Erlangen, Germany) ($n = 16$):

1. 3D T1-weighted magnetisation prepared – rapid gradient echo sequence (MPRAGE, field of view (FoV) = 256 mm, matrix 256 × 256, 176 slices, slice thickness 1 mm, voxel dimension 1.0 × 1.0 × 1.0 $mm^3$, repetition time (TR) = 1900 ms, echo time (TE) = 2.26 ms, inversion time (TI) = 900 ms, flip-angle = 9°, bandwidth (BW) = 200 Hz/Pixel, parallel imaging (GRAPPA) with factor 2).
2. Diffusion-weighted imaging (FoV = 256 mm, matrix 128 × 128, slice thickness 2 mm, distance factor 0%, voxel dimension 2.0 × 2.0 × 2.0 $mm^3$, TR = 7900 ms, TE = 90 ms, BW = 1502 Hz/Pixel, 42 diffusion encoding gradients, three intermittent non-weighted b0 images ($b = 0$ s/$mm^2$), high b-value $b = 1000$ s/$mm^2$, GRAPPA with factor 2)

1.5 T MRI system (Avanto, Siemens Healthineers, Erlangen, Germany) ($n = 3$):

1. 3D T1-weighted magnetization prepared – rapid gradient echo sequence (MPRAGE, field of view (FoV) = 256 mm, matrix 246 × 256, 176 slices, slice thickness 1 mm, voxel dimension 1.0 × 1.0 × 1.0 $mm^3$, repetition-time (TR) = 2090 ms, echo-time (TE) = 3.08 ms, inversion time (TI) = 1100 ms, flip-angle = 15°, bandwidth (BW) = 130 Hz/Pixel, parallel imaging (GRAPPA) with factor 2)
2. Diffusion-weighted imaging (FoV = 256 mm, matrix 128 × 128, slice thickness 2 mm, distance factor 0%, voxel dimension 2.0 × 2.0 × 2.0 $mm^3$, TR = 7500 ms, TE = 93 ms, BW = 1347 Hz/Pixel, 30 diffusion encoding gradients, three intermittent non-weighted b0 images ($b = 0$ s/$mm^2$), high b-value $b = 1000$ s/$mm^2$, GRAPPA with factor 2).

1.5 T MRI system (Espree, Siemens Healthineers, Erlangen, Germany) ($n = 1$):

1. 3D T1-weighted magnetisation prepared – rapid gradient echo sequence (MPRAGE, field of view (FoV) = 256 mm, matrix 246 × 256, 176 slices, slice thickness 1 mm, voxel dimension 1.0 × 1.0 × 1.0 $mm^3$, repetition-time (TR) = 2090 ms, echo-time (TE) = 3.08 ms, inversion time (TI) = 1100 ms, flip-angle = 15°, bandwidth (BW) = 130 Hz/Pixel, parallel imaging (GRAPPA) with factor 2)
2. Diffusion-weighted imaging (FoV = 256 mm, matrix 128 × 128, slice thickness 2 mm, distance factor 0%, voxel dimension 2.0 × 2.0 × 2.0 $mm^3$, TR = 10900 ms, TE = 105 ms, BW = 1221 Hz/Pixel, 30 diffusion encoding gradients, three intermittent non-weighted b0 images (b = 0 $s/mm^2$), high b-value b = 1000 $s/mm^2$, GRAPPA with factor 2)

#### Supplementary Figures

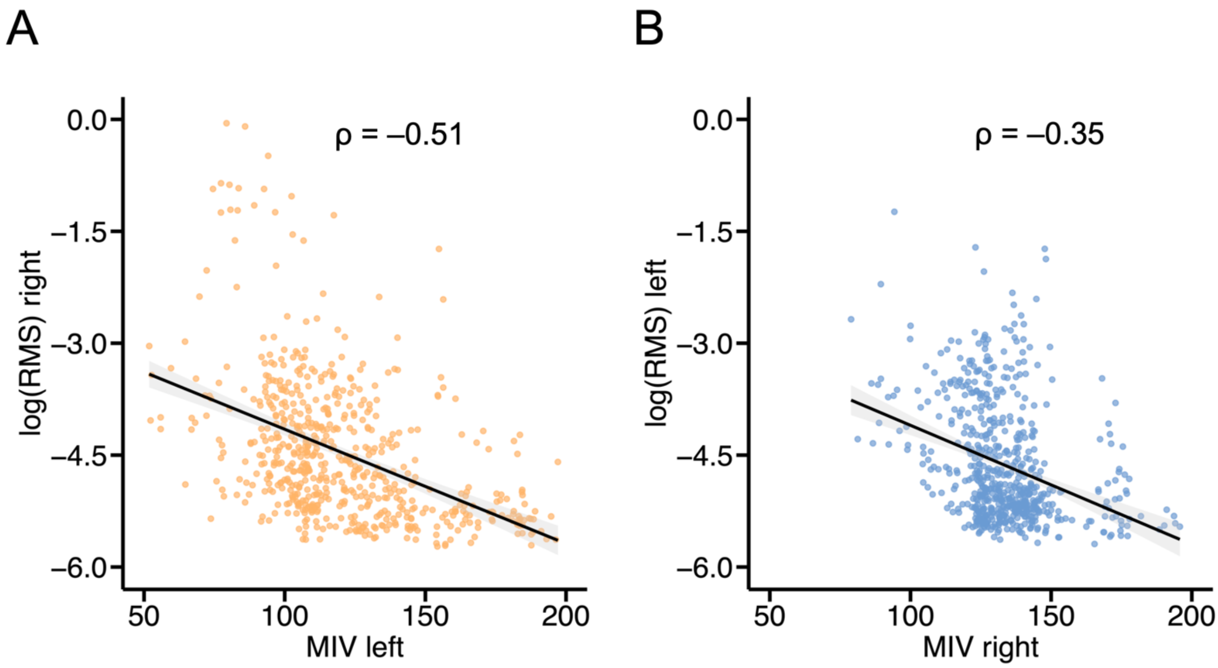


**Supplementary Figure 1 Local stimulation effects.** Scatter plots show Spearman correlations between mean intensity values (MIV) and contralateral root mean square (RMS) scores for the left (**A**) and right (**B**) hemisphere. Black lines indicate linear regression fits.

### Supplementary Tables

**Supplementary Table 1 Top seven loadings per hemisphere for component 1.**

| Hemisphere | Region | Loading Value |
|---|---|---|
| Left | Pallidum | 0.231 |
| | Putamen | 0.228 |
| | Caudate | 0.221 |
| | Thalamus-Proper | 0.220 |
| | Cerebellum-Cortex | 0.217 |
| | precentral | 0.216 |
| | postcentral | 0.207 |
| Right | Caudate | 0.112 |
| | precentral | 0.110 |
| | Pallidum | 0.108 |
| | postcentral | 0.102 |
| | Thalamus-Proper | 0.096 |
| | Putamen | 0.095 |
| | frontalpole | 0.082 |

**Supplementary Table 2 Top seven loadings per hemisphere for component 2.**

| Hemisphere | Region | Loading Value |
|---|---|---|
| Left | entorhinal | 0.220 |
| | superiorfrontal | 0.171 |
| | parsopercularis | 0.165 |
| | pericalcarine | 0.152 |
| | temporalpole | 0.142 |
| | Putamen | 0.140 |
| | paracentral | 0.135 |
| Right | middletemporal | 0.436 |
| | rostralmiddlefrontal | 0.424 |
| | inferiorparietal | 0.417 |
| | frontalpole | 0.400 |
| | supramarginal | 0.393 |
| | lateralorbitofrontal | 0.389 |
| | Hippocampus | 0.387 |

**Supplementary Table 3 Top seven loadings per hemisphere for component 3.**

| Hemisphere | Region | Loading Value |
|---|---|---|
| Left | posteriorcingulate | 0.148 |
| | caudalanteriorcingulate | 0.140 |
| | paracentral | 0.131 |
| | rostralanteriorcingulate | 0.109 |
| | isthmuscingulate | 0.087 |
| | precuneus | 0.086 |
| | Hippocampus | 0.083 |
| Right | superiorfrontal | 0.236 |
| | Pallidum | 0.230 |
| | Cerebellum-Cortex | 0.229 |
| | Putamen | 0.229 |
| | Thalamus-Proper | 0.227 |
| | precentral | 0.225 |
| | paracentral | 0.223 |

**Supplementary Table 4 Top seven loadings per hemisphere for component 4.**

| Hemisphere | Region | Loading Value |
|---|---|---|
| Left | middletemporal | 0.197 |
| | inferiortemporal | 0.181 |
| | lateraloccipital | 0.178 |
| | supramarginal | 0.170 |
| | lingual | 0.168 |
| | superiortemporal | 0.162 |
| | inferiorparietal | 0.152 |
| Right | Pallidum | 0.258 |
| | Cerebellum-Cortex | 0.240 |
| | superiorfrontal | 0.239 |
| | Putamen | 0.229 |
| | rostralanteriorcingulate | 0.225 |
| | lingual | 0.225 |
| | precentral | 0.222 |

**Supplementary Table 5 Top seven loadings per hemisphere for component 5.**

| Hemisphere | Region | Loading Value |
|---|---|---|
| Left | medialorbitofrontal | 0.427 |
| | rostralanteriorcingulate | 0.379 |
| | insula | 0.299 |
| | Hippocampus | 0.296 |
| | superiortemporal | 0.281 |
| | frontalpole | 0.264 |
| | rostralmiddlefrontal | 0.255 |
| Right | rostralanteriorcingulate | 0.243 |
| | Cerebellum-Cortex | 0.216 |
| | parsorbitalis | 0.205 |
| | paracentral | 0.201 |
| | caudalanteriorcingulate | 0.184 |
| | superiorfrontal | 0.157 |
| | supramarginal | 0.136 |

**Supplementary Table 6 Top seven loadings per hemisphere for component 6.**

| Hemisphere | Region | Loading Value |
|---|---|---|
| Left | superiorfrontal | 0.294 |
| | transversetemporal | 0.273 |
| | caudalanteriorcingulate | 0.266 |
| | lateraloccipital | 0.240 |
| | superiortemporal | 0.221 |
| | lingual | 0.212 |
| | supramarginal | 0.200 |
| Right | parsopercularis | 0.348 |
| | parstriangularis | 0.277 |
| | fusiform | 0.266 |
| | superiorfrontal | 0.242 |
| | temporalpole | 0.241 |
| | entorhinal | 0.236 |
| | cuneus | 0.216 |
